# B cell response six months after SARS-CoV-2 mRNA vaccination in people living with HIV under antiretroviral therapy

**DOI:** 10.1101/2022.07.01.22277132

**Authors:** Jacopo Polvere, Massimiliano Fabbiani, Gabiria Pastore, Ilaria Rancan, Barbara Rossetti, Miriam Durante, Sara Zirpoli, Enrico Morelli, Elena Pettini, Simone Lucchesi, Fabio Fiorino, Mario Tumbarello, Annalisa Ciabattini, Francesca Montagnani, Donata Medaglini

## Abstract

**Background:** SARS-CoV-2 mRNA vaccines have demonstrated high immunogenicity in healthy subjects and preliminary results for people living with HIV (PLWHIV) are promising too. We have previously reported the persistence of spike-specific circulating IgG and memory B cells in healthy adults up to six months after mRNA SARS-CoV-2 vaccination. Unfortunately, limited longitudinal data are available for PLWHIV and no evidence of persistent spike-specific B cells have been reported yet.

**Methods:** We investigated the humoral response and the persistence of spike-specific memory B cells up to six months after vaccination with two doses of mRNA vaccines in 84 PLWHIV under ART and compared them to healthy controls (HCs). Humoral response was analyzed with enzyme-linked immunosorbent assay and with an angiotensin-converting enzyme 2 (ACE2) and receptor binding domain (RBD) inhibition assay. PBMCs were analyzed with a cytofluorimetric approach for B cell phenotyping.

**Findings:** Spike-specific IgG peaked 1 month after second dose and persisted up to six months after vaccination with no significant differences compared to HCs. The stratification of patients according to CD4+ T cell count showed a significantly lower IgG response in case of CD4<350/µl, remarking the relevance of immune reconstitution. The ability of IgG of blocking the binding between ACE2 and RBD was detected in 58·4% of PLWHIV, compared to 86·2% in HCs. The amount of circulating spike-specific memory B cells detected in PLWHIV six months after vaccination was not significantly different from HCs, while there was prevalence of antigen-specific double negative (IgD-/CD27-) cells, compared to controls.

**Interpretation:** In conclusion, the majority of PLWHIV developed spike-specific humoral and B cell responses that persist for at least six months after SARS-CoV-2 mRNA vaccination. However, hints of HIV-dependent immune impairment were revealed by altered spike-specific B cell phenotypes and by reduced spike-specific humoral response in patients with low CD4+ T cell count (<350/µl).

## Introduction

People affected by immunodeficiency conditions, such as HIV infection, are known to be more susceptible to infections but the effect of impaired immunity in SARS-CoV-2 infection and COVID-19 progression is still hard to be stated (1). There is no consensus on the role of chronic HIV infection in COVID-19 outcome, since in some cohorts it appears as a negligible risk factor but other studies highlight the higher rate of hospitalization and death in PLWHIV, especially for patients with low CD4+ T cells count and with detectable viremia (2–4). From an immunological point of view, the CD4+ T cells/µl count and the CD4+/CD8+ T cell ratio, as well the antiretroviral regimens (ART) and compliance, are relevant factors to mount an effective immune response in PLWHIV (5,6). A recent study demonstrated that SARS-CoV-2 infected PLWHIV under effective ART, with a good immunological recovery and adequate virological control, can develop immune responses that are similar to healthy controls, both at humoral and T cell level (7).

Differently from other immunocompromised cohorts, most PLWHIV present high immune responsiveness for SARS-CoV-2 mRNA vaccine platforms (8,9), yet the anti-Spike immunity can even be absent in PLWHIV with profound immune dysfunction vaccinated with two doses (10).

As a consequence of waning immune protection against SARS-CoV-2 infection and the emerging viral variants of concerns, the FDA have recently approved a fourth dose with mRNA vaccines for immunocompromised (11). Whether or when to globally plan other future booster doses in PLWHIV, is a burning question. Immunological studies on healthy subjects vaccinated with nanoparticles-based mRNA formulations (12) clearly show the persistence of circulating Spike-specific antibodies and immune memory cells at least after six months from vaccination (13,14). Up to date, longitudinal data regarding SARS-CoV-2 vaccine humoral response in PLWHIV do not suggest a reduced persistence of circulating antibody compared to healthy subjects (15–17). However, considering HIV-induced immunological impairments (18), It is of primary importance to investigate the induction and persistence of spike-specific memory B cells in PLWHIV upon vaccination. Indeed, there are no published studies that address the memory B cell compartment, which is considered essential to guarantee a long-term immunity and reactivation at the antigen encounter (13).

In this longitudinal study, we evaluated the B cell response and the kinetic of the humoral response against SARS-CoV-2 for six months after the mRNA vaccination in PLWHIV, that are currently on ART, and we compared data with healthy controls.

## Methods

### Study cohort

Plasma and peripheral blood mononuclear cells (PBMC) samples were obtained from adult PLWHIV under different ART regimens and from healthy controls (HCs), who received two doses of mRNA vaccines, either mRNA-1273 (Moderna) or BNT162b2 (Pfizer), 3 or 4 weeks apart according to national schedules. History of SARS-CoV-2 infection before or after vaccination were reasons for exclusion. All participants provided written informed consent before participation to the study. Study participants were recruited at the Infectious and Tropical Diseases Unit, Azienda Ospedaliera Universitaria Senese (Siena, Italy). The study was performed in compliance with all relevant ethical regulations and the protocol was approved by local Ethical Committee for Clinical experimentation of Regione Toscana Area Vasta Sud Est (CEASVE), protocol code 19479 PATOVAC v1.0 of 03 Mar 2021, approved on 15 Mar 2021 and protocol code 18869 IMMUNO_COV v1.0 of 18 Nov 2020, approved on 21 Dec 2020 for HCs.

### Plasma and Peripheral Blood Mononuclear Cells Isolation

Venous blood samples were collected in heparin-coated blood tubes (BD Vacutainer) at baseline (pre v1), before the second dose (pre v2), and 1, 2 and 5 months post the second dose (+30 v2, +60 v2 and +150 v2, respectively. PBMCs were isolated by density-gradient sedimentation, using Ficoll-Paque (Lymphoprep, Meda, Italy). Isolated PBMC were then cryopreserved in a cell recovery medium [10% DMSO (Thermo Fisher Scientific) and 90% heat inactivated fetal bovine serum (Sigma Aldrich)] and stored in liquid nitrogen until used. Plasma samples were stored at -80°C.

### ELISA

Humoral spike-specific IgG were tested on Maxisorp microtiter plates (Nunc, Denmark) coated with recombinant SARS-CoV-2 Spike S1+S2 ECD (1 μg/ml protein; Sino Biological), as previously reported (19). Briefly, plates were blocked (PBS and 5% skimmed milk powder, 0.05% Tween 20) and the added with heat-inactivated plasma samples and titrated in twofold dilutions in duplicate. Anti-human horseradish peroxidase (HRP)-conjugated IgG was added in diluent buffer for 1 h at RT, and then plates were developed with 3,3’,5,5’-Tetramethylbenzidine (TMB; Thermo Fisher Scientific) substrate for 10 min at RT, followed by addition of 1M stop solution. Absorbance at 450 nm was measured on Multiskan FC Microplate Photometer (Thermo Fisher Scientific).

### ACE2/RBD inhibition assay

ACE2/RBD inhibition was tested with a SARS-CoV-2 surrogate virus neutralization test (sVNT) kit (cPass™, Genscript), according to the manufacturer protocol and as previously reported (13). Results are reported as follows: percentage inhibition = (1 - sample OD value/negative control OD value) * 100. Inhibition values ≥30% are regarded as positive results, while values <30% as negative results, as indicated by the manufacturer.

### Multiparametric flow cytometry

Two million of PBMCs incubated with BD human FC block (BD Biosciences) and stained with biotinylated spike S1+S2 ECD-His recombinant biotinylated-protein (Sino Biological) conjugated with SA-R-Phycoerythrin (PE) and RBD recombinant biotinylated-protein (BioLegend) conjugated with SA-Allophycocyanin (APC), together with the following fluorescent antibodies: CD3-BV650 (clone OKT3); CD21-FITC (clone B-LY4), CD19-BUV395 (clone SJ25C1), CD10-PECF594 (clone HI10A), IgM-BV605 (clone G20-127), IgD-BV711 (clone IA6-2), CD27-BV786 (clone O323), CD11c-BB700 (clone 3.9), CD20-APCH7 (clone 2H7), CD38-BUV737 (clone HB7), IgG-PE-Cy7 (clone G18-145, all from Becton Dickinson), IgA-Vio blue (clone IS11-8E10, Miltenyi Biotec). All antibodies were titrated for optimal dilution. Following surface staining, cells were washed once with PBS and labeled with Zombie Aqua Fixable Viability Kit (Thermofisher) according to the manufacturer instruction. Cells were fixed in BD fixation solution (BD Biosciences) and acquired with SO LSRFortessa X20 flow cytometer (BD Biosciences). Data analysis was performed using FlowJo v10 (TreeStar, USA).

### t-SNE visualization

The B cell population analyzed in our data set was gated as live, singlet, CD3−/CD19+ cells using FlowJo v10 (TreeStar, USA) while antigen specific B cells were gated as Spike+/RBD+ cells. B cell files were then exported as .fcs file and imported in R environment as flowSet object, that was then compensated with FlowCore package 2.6.0 and logicle transformed (20). t-Distributed Stochastic Neighbor Embedding (t-SNE) dimensionality reduction (21) was performed with Rtsne package v0.15. Expression values of each marker were normalized as z-scores (mean=0 and standard deviation=1). An equal number of cells (25000) were downsampled from PLWHIV and healthy donor samples, then Rtsne function was run setting perplexity = 100, selected as optimal parameter value in a range between 5 and 200. B cells were analyzed with manual gating and labels of different B cell populations were imported in R environment using GetFlowJoLabels function from FlowSOM package (v2.2.0) and visualized with function Contour from FlowViz package (v1.58.0).

### Statistical Analysis

Descriptive statistics [number, proportion, median, interquartile range (IQR), 95% confidence intervals (CI)] were used to describe the baseline characteristics of patients. Categorical variables were compared between groups using the Chi-square test or Fisher’s exact test, as appropriate. Continuous variables were compared using the non-parametric Mann-Whitney U test. The Kruskal–Wallis test, followed by Dunn’s post-test for multiple comparisons, was used to assess the statistical differences of ELISA titers and B cell subpopulations among different groups of subjects for each time point. Fisher’s exact test was used to assess the statistical differences in percentages of ACE2/RBD inhibition positive subjects between groups. Clinical and laboratory variables of PLWHIV, potentially associated with log-transformed ELISA titers, ACE2/RBD inhibition percentage and with inhibition value ≥30% were investigated by linear or logistic regression models, as appropriate. Factors associated with the dependent variables at univariate analysis, together with absolute CD4+ T cells count, CD4+ T cells percentage and CD4+/CD8+ ratio, were include in a multivariate model. A *p*-value ≤ 0.05 was considered significant. Analyses were performed using GraphPad Prism v9 (GraphPad Software, San Diego, CA, USA) and SPSS Software, version 23.0 (SPSS Inc., Chicago, IL).

### Role of the funding source

The sponsor of the study had no role in study design, data collection, data analysis, data interpretation, or writing of the report. The sponsor approved the decision to submit the paper for publication.

## Results

A total of 84 PLWHIV with a median age of 52 years (IQR 46-58) were enrolled in the study, of whom 64 (76.2%) were male and 20 (23.8%) were female. Among them, 57 (67.9%) had >500 CD4+ T cells/µl (High CD4+), 13 (15.5%) were in the 350-500 CD4+ T cells/µl range (Average CD4+) and 14 (16.7%) had values <350 CD4+ T cells/µl (Low CD4+). At baseline, 28 (33.3%) PLWHIV showed optimal immunological recovery, defined as CD4+ T cells count ≥500 cell/µl plus CD4+ T cells % ≥ 30% plus CD4/CD8 ratio ≥ 1 (22,23). Clinical and demographic data of PLWHIV cohorts were collected before the first dose for the statistical analysis and reported in Supplementary table 1. No changes in treatment or significant variation in CD4+ T cells count were observed during the study. The HCs group was composed of 79 healthy volunteers with a median age of 52 years (IQR 45-60) of whom 22 (27.8%) were males and 57 (72.2%) were females. A slightly higher body mass index (BMI) (25.1 vs 23.7, p=0.037) was observed in PLWHIV. Moreover, HCs were more frequently vaccinated with BNT162b2 vaccine when compared to PLWHIV who mostly received mRNA-1273 (87.3% vs 48.8%, p<0.001.)

### Spike-specific antibody response following SARS-CoV-2 mRNA vaccination in PLWHIV

Spike-specific IgG induced by SARS-CoV-2 vaccination were assessed by ELISA on plasma samples collected from PLWHIV at different time points (figure 1). Anti-spike IgG significantly increased after the first dose, reaching a geometric mean titer (GMT) of 2105 at pre v2 [95% CI 1514 to 2928; titers range 320–20480; P≤0.001 vs. pre v1; Figure 2A]. Antibody titres increased again after the second dose, reaching a peak at +30 v2, with a GMT of 14482 (95% CI 10531 to 19915; titers range 640–81920; P≤0.001 vs. pre v1 and pre v2). Significant antibody titers were still observed 2 months post boost (+60 v2) (GMT 6684; 95% CI 5086 to 8785; titers range 320-40960; P≤0.001 vs. pre v1 and P≤0.01 vs. pre v2) and 5 months post boost (+150 v2) (GMT 4974; 95% CI 3218 to 7690; titers range 320-20480; P≤0.001 vs. pre v1) despite a progressive physiological decline over time. No statistically significant differences were observed between 2 months (+60 v2) and 5 months (+150 v2) post-boost (P>0.99), as well as between GMT at pre-boost (pre v2) and at 5 months post-boost (+150 v2) (P=0.46).

**Figure 1.**
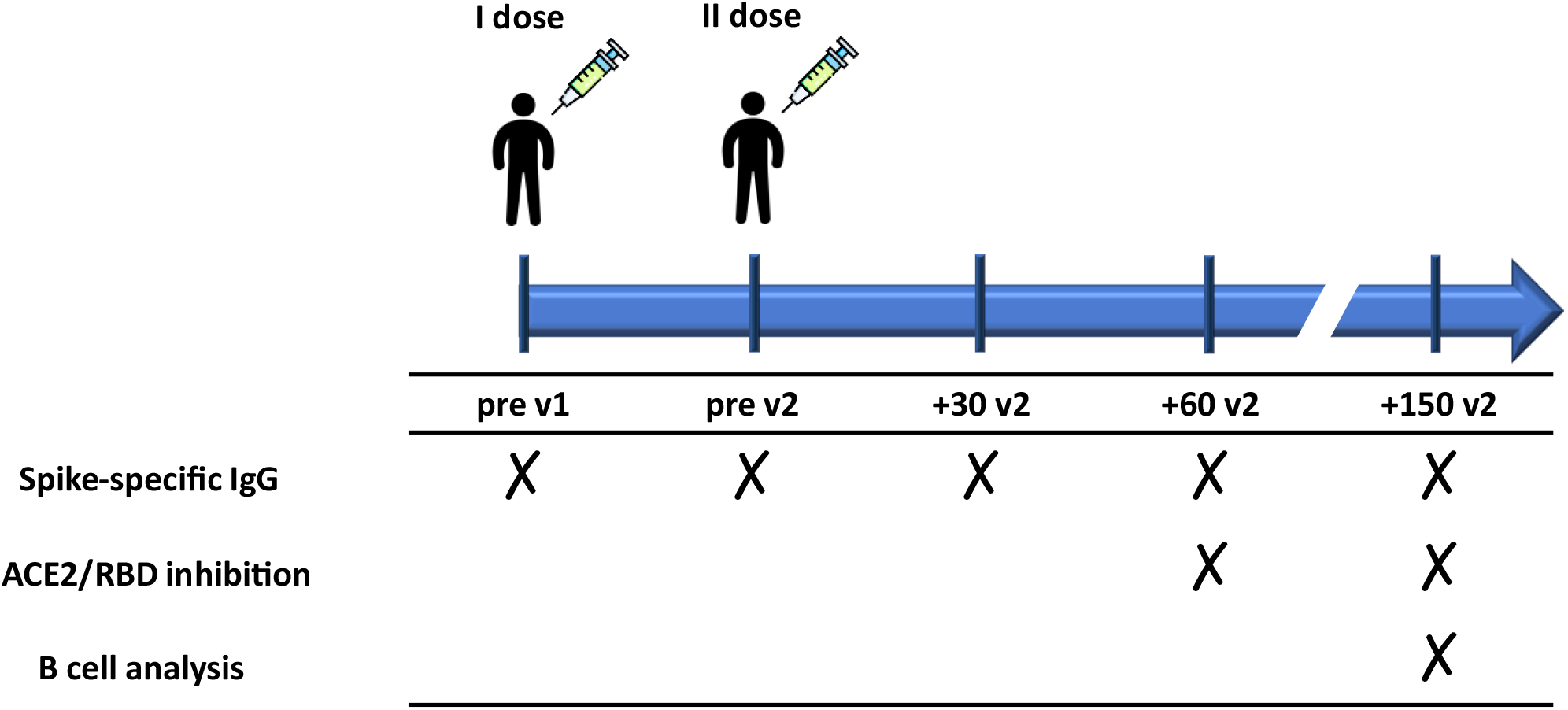
Schematic representation of the study design. PLWHIV (84 subjects) and healthy controls (HCs, 79 subjects) vaccinated with two doses of a SARS-CoV-2 mRNA vaccine (BNT162b2 Pfizer-BioNTech or mRNA-1273 Moderna) 3-4 weeks apart, respectively, were enrolled in the study. Blood samples were collected at pre v1 (day 0, baseline), pre v2, +30 v2 (1 month post second dose), +60 v2 (2 months post second dose), and +150 v2 (5 months post second dose). Plasma samples were tested for spike-specific IgG and ACE2/RBD inhibition while peripheral blood mononuclear cells (PBMCs) were analyzed for spike-specific B cell response.

**Figure 2.**
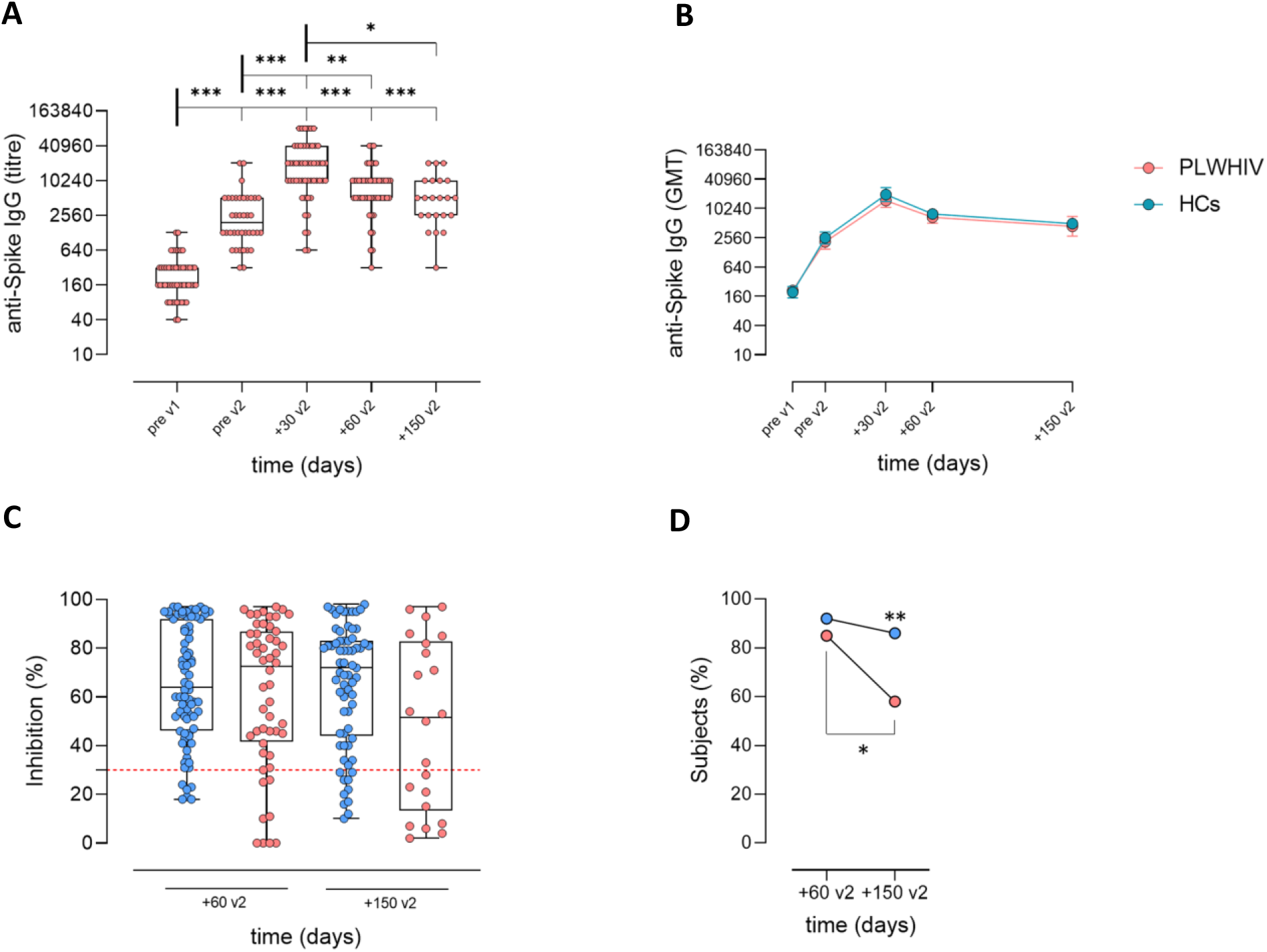
Spike-specific antibody response following SARS-CoV-2 mRNA vaccination detected in plasma. A) IgG anti-Spike detected at pre v1, pre v2, +30 v2, +60 v2 and +150 v2 in PLWHIV. Data are presented as box and whiskers diagram showing the minimum and maximum of all the data. Kruskal-Wallis test, followed by Dunn’s post test for multiple comparisons, was used for assessing statistical differences between groups. *P≤ 0.05; **P ≤ 0.01; ***P≤ 0.001. B) IgG titers comparison between HCs and PLWHIV. Antibody titres are expressed as the reciprocal of the dilution of sample reporting a double OD value compared to the background. Data are presented as geometric mean titers (GMT) with error. Kruskal-Wallis test was used for assessing statistical differences between groups. C, D) Surrogate virus neutralization assay (sVN) performed at +60 v2 and +150 v2. C) Data are reported as ACE2/RBD binding inhibition percentage with box and whiskers diagram showing the minimum and maximum of all the data. A threshold (dotted red line) was placed at 30% inhibition percentage to discriminate between positive and negative samples. Kruskal-Wallis test, followed by Dunn’s post-test for multiple comparisons, was used for assessing statistical differences between groups. D) Data are reported as percentage of positive subjects over all samples tested at each time point. Fisher’s Exact Test was used to assessing statistical differences between groups. *P≤ 0.05; **P≤ 0.01.

The curve of spike-specific IgG observed in PLWHIV perfectly overlaid to the one measured in HC (figure 2B) Both cohorts reached their maximum titers after the second dose (HCs GMT at +7 v2: 19677; 95% CI 14183 to 27300) and their response progressively declined over time. No statistically significant differences were observed between the two groups for any time point of the study.

A stratification based on CD4+ T cell/µl (≤350 Low CD4+, 350-500 Average CD4+ and ≥500 High CD4+) was done to highlight a possible predictive value for the humoral response (figure S1). The statistical analysis revealed a significant difference in antibody response at +30 v2 between the High CD4+ (GMT 19038; 95% CI 14328 to 25297) and Low CD4+ (GMT 1612; 95% CI 539 to 4821) (P≤0.001) and between Average CD4+ (GMT 22119; 95% CI 11248 to 43499) and Low CD4+ (P≤0.001; figure S1). Pearson test was used to assess the correlation between absolute CD4+ T cells/µl and ELISA IgG titers. A significant positive correlation was found at pre v2 (r=0.4421; P≤0.01), +30 v2 (r=0.4473; P≤0.001) and at +60 v2 (r=0.2987; P≤0.05) but not at +150 v2.

We also investigate HIV-related variables associated to IgG titers at +30 v2 by linear regression analysis (Table 1). At multivariate analysis, patients with <350 cells/μl confirmed to elicit lower IgG titers (mean change -1.02, 95% CI -1.39 to -0.65, p<0.001 when compared to those with CD4+>350 cells/μl) after adjusting for having CD4+ T cells % ≥ 30% and CD4+/CD8+ ratio ≥1.

**Tables 1.** Variables associated to log-transformed IgG titer at 1 month post boost dose (+30 v2) (univariable and multivariable linear regression analysis).

|  | Univariable analysis |  | Multivariable analysis |  |
| --- | --- | --- | --- | --- |
|  | Mean change<br>(95% CI) | p | Adjusted mean change<br>(95% CI) | p |
| Age, per +10 years | 0.11 (-0.05 / 0.26) | 0.169 |  |  |
| Male gender | -0.10 (-0.43 / 0.23) | 0.563 |  |  |
| Type of vaccine (mRNA-1273 vs BNT162b2) | -0.07 (-0.37 / 0.22) | 0.623 |  |  |
| BMI, Kg/m<sup>2</sup> | -0.00 (-0.03 / 0.02) | 0.836 |  |  |
| IDU | 0.03 (-0.36 / 0.43) | 0.864 |  |  |
| Years from HIV infection, per +10 years | 0.03 (-0.12 / 0.17) | 0.731 |  |  |
| CDC stage C | -0.15 (-0.53 / 0.23) | 0.426 |  |  |
| HBV or HCV coinfection | -0.05 (-0.36 / 0.25) | 0.736 |  |  |
| Zenith HIV-RNA, per +1 log copies/mL | -0.01 (-0.28 / 0.27) | 0.946 |  |  |
| CD4 cell count at nadir, per +100 cell/mm<sup>3</sup> | 0.07 (-0.02 / 0.16) | 0.124 |  |  |
| Years from first ART, per +10 years | 0.06 (-0.12 / 0.23) | 0.526 |  |  |
| InSTI + 2NRTI | -0.10 (-0.40 / 0.20) | 0.503 |  |  |
| Baseline HIV-RNA <50 cp/mL | 0.25 (-0.28 / 0.79) | 0.347 |  |  |
| Time from last HIV-RNA >50 copies/mL, per +10 years | -0.02 (-0.47 / 0.44) | 0.942 |  |  |
| CD4 cell count at baseline, per +100 cell/mm<sup>3</sup> | 0.04 (-0.01 / 0.08) | 0.102 |  |  |
| CD4 cell count at baseline $\leq 350$ cell/mm<sup>3</sup> | -1.08 (-1.41 / -0.75) | **<0.001** | -1.02 (-1.39 / -0.65) | **<0.001** |
| CD4% $\geq 30\%$ | 0.36 (0.09 / 0.64) | **0.009** | 0.01 (-0.30 / 0.32) | 0.947 |
| CD4/CD8 ratio $\geq 1$ | 0.33 (0.06 / 0.60) | **0.019** | 0.11 / -0.19 / 0.40) | 0.459 |

**Abbreviations:** ART, antiretroviral therapy; BMI, body mass index; CI, confidence intervals; HBV, hepatitis B virus; HCV, hepatitis C virus; IDU, injecting drug users; InSTI, integrase strand transfer inhibitors; MSM, men who have sex with men; NRTI, nucleoside reverse transcriptase inhibitors; NNRTI, non nucleoside reverse transcriptase inhibitors; OIR, optimal immunological recovery; PI, protease inhibitors.

A surrogate virus neutralization assay was employed to evaluate the persistence of the ability of vaccine-induced antibodies to block the ACE2/RBD interaction at +60 v2 and +150 v2. Results were reported as percentages of ACE2/RBD inhibition (figure 2C) and as percentage of patients with inhibition values ≥ 30% (figure 2D). PLWHIV cohort registered a significant reduction in the percentage of patients with a positive value of inhibiting antibodies overtime (84.6% at +60 v2 vs 58.4% at +150 v2; P≤0.05, figure 2D), while the HCs group, maintained a stable value (92.1% at +60 v2 vs 86.2% at +150 v2; P>0.9999). The frequency of inhibition positive PLWHIV at 5 months post-second dose was significantly lower compared to HCs (58.4% vs 86.2%, respectively; P≤0.01).

We also investigated HIV-related variables associated to ACE2/RBD inhibition percentage at 2 months post second dose (+60 v2) by linear regression analysis (Supplementary table 2). A trend toward an association between having CD4+ T cell count at baseline ≤350 cell/µl and a lower inhibition percentage was observed at univariate analysis (mean change -19.30%, 95% CI -40.99 / 2.40, P=0.080), but this was not confirmed after adjusting for having CD4+ % ≥30% and CD4/CD8 ratio ≥1.

Moreover, we explored variables associated to the percentage of inhibition positive PLWHIV at 2 months post boost dose (+60 v2) by logistic regression analysis (Table 2). At multivariate analysis, a higher CD4+ % was associated with a higher probability to obtain inhibition (adjusted odd ratio 1.12, 95% CI 1.00-1.25, P=0.047), after adjusting for absolute CD4+ count and CD4/CD8 ratio.

**Tables 2.** Variables associated to the percentage of inhibition positive PLWHIV at 2 months post boost dose (+60 v2) (univariable and multivariable logistic regression analysis).

|  | Univariable analysis |  | Multivariable analysis |  |
| --- | --- | --- | --- | --- |
|  | OR<br>(95% CI) | p | Adjusted OR<br>(95% CI) | p |
| Age, per +10 years | 1.36 (0.67-2.79) | 0.395 |  |  |
| Male gender | 1.00 (0.18-5.70) | 1.000 |  |  |
| Type of vaccine (mRNA-1273 vs BNT162b2) | 1.43 (0.30-6.88) | 0.655 |  |  |
| BMI, Kg/m<sup>2</sup> | 1.07 (0.84-1.37) | 0.577 |  |  |
| IDU | 0.90 (0.09-8.89) | 0.926 |  |  |
| Years from HIV infection, per +10 years | 1.09 (0.52-2.26) | 0.819 |  |  |
| CDC stage C | 0.32 (0.06-1.63) | 0.169 |  |  |
| HBV or HCV coinfection | 2.06 (0.23-18.78) | 0.522 |  |  |
| Zenith HIV-RNA, per +1 log copies/mL | 0.46 (0.10-2.10) | 0.315 |  |  |
| CD4 cell count at nadir, per +100 cell/mm<sup>3</sup> | 1.20 (0.74-1.93) | 0.450 |  |  |
| Years from first ART, per +10 years | 1.73 (0.46-6.51) | 0.418 |  |  |
| Type of ART: |  |  |  |  |
| -InSTI + 2NRTI | Ref |  |  |  |
| -PI + 2NRTI | 1.56 (0.15-16.72) | 0.712 |  |  |
| -NNRTI + 2NRTI | 1.40 (0.23-8.78) | 0.715 |  |  |
| Baseline HIV-RNA <50 cp/mL | 1.95 (0.18-21.54) | 0.585 |  |  |
| Time from last HIV-RNA >50 copies/mL, per +10 years | 4.27 (0.24-76.54) | 0.325 |  |  |
| CD4 cell count at baseline, per +100 cell/mm<sup>3</sup> | 0.85 (0.66-1.09) | 0.193 | 0.89 (0.67-1.17) | 0.389 |
| CD4 cell count at baseline ≤350 cell/mm<sup>3</sup> | 0.26 (0.05-1.40) | 0.117 |  |  |
| CD4%, per +1% increase | 1.09 (0.01-1.18) | **0.029** | 1.12 (1.00-1.25) | **0.047** |
| CD4% ≥30% | 2.41 (0.51-11.37) | 0.267 |  |  |
| CD4/CD8 ratio, per +1 increase | 1.99 (0.45-8.85) | 0.369 | 0.55 (0.14-2.13) | 0.387 |
| CD4/CD8 ratio ≥1 | 2.08 (0.38-11.48) | 0.402 |  |  |

**Abbreviations:** ART, antiretroviral therapy; BMI, body mass index; CI, confidence intervals; HBV, hepatitis B virus; HCV, hepatitis C virus; IDU, injecting drug users; InSTI, integrase strand transfer inhibitors; MSM, men who have sex with men; NRTI, nucleoside reverse transcriptase inhibitors; NNRTI, non nucleoside reverse transcriptase inhibitors; OIR, optimal immunological recovery; PI, protease inhibitors.

### Spike-specific memory B cells following SARS-CoV-2 mRNA vaccination in PLWHIV

The immune memory induced by SARS-CoV-2 vaccination was evaluated on PBMCs samples collected at +150 v2, by multiparametric flow cytometry (Figure 3). Circulating spike-specific cells were identified using fluorescent tetramerized full-spike protein (S) and receptor binding domain (RBD). Spike-specific (S+RBD+) B cells were identified applying the gating strategy reported in Figure 3A. The mean values of S^+^RBD^+^ B cells observed in blood of PLWHIV patients at +150 v2 were not significantly different with HCs (0.343 ± 0.547% vs 0.171 ± 0.179%; P=0.9345) even though 42% of subjects PLWHIV showed a very low amount of spike-specific B cells (<0.1 % over all B cells) (Figure 3B).

**Figure 3.**
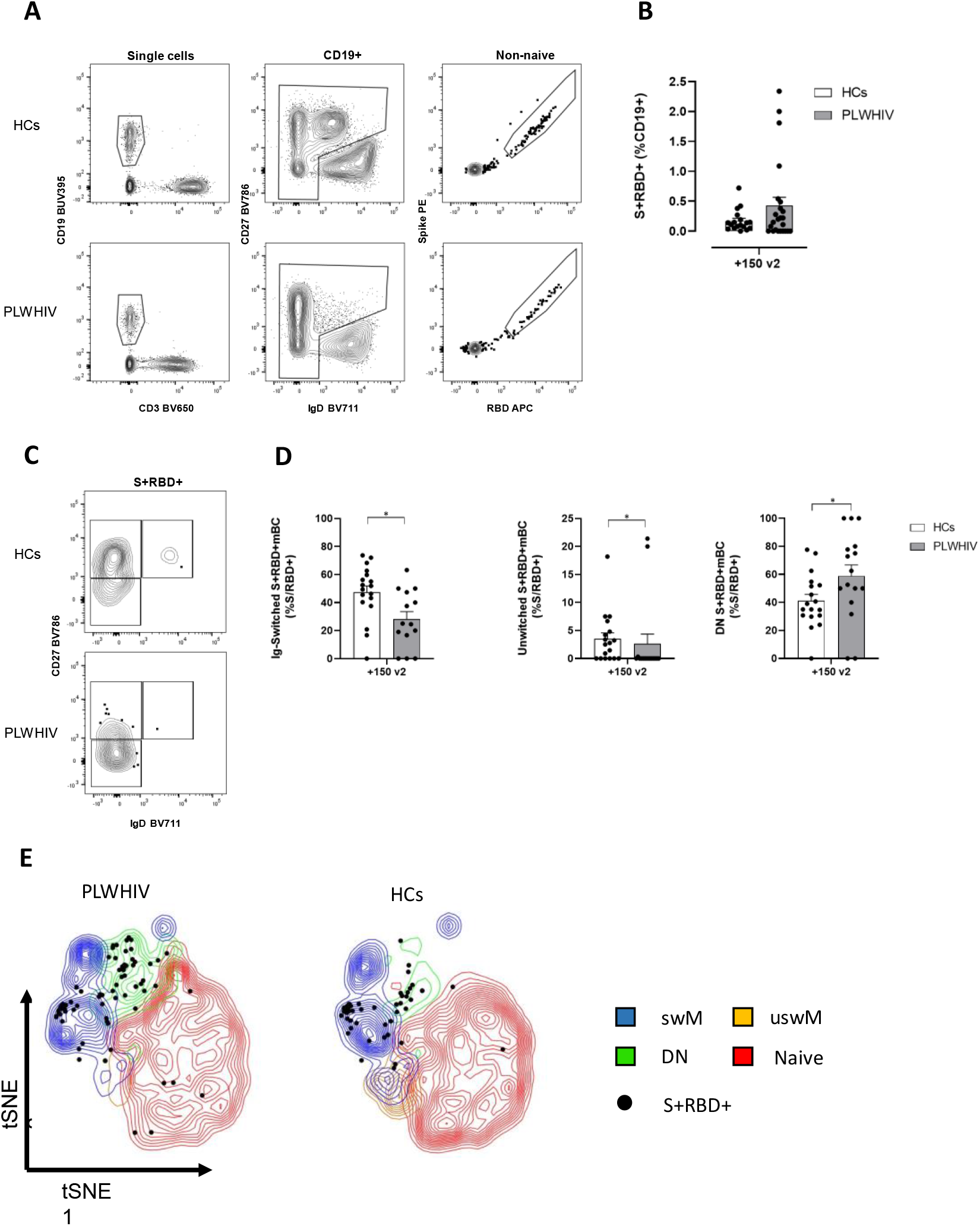
Analysis of Spike-specific memory B cells at 6 months after the beginning of the vaccination cycle (+150 v2). A) Gating strategy applied for identifying non-naïve Spike-specific B cells (S+/RBD+) in PLWHIV and HCs. B) Percentages of S+RBD+ cells identified within CD19+ cells pool in each group. C) Contour plot analysis of CD27 versus IgD within non-naïve S+RBD+ B cells, for identifying Ig-switched (IgD-/CD27+), Unswitched (IgD+/CD27+) and Double negative (DN)(IgD-/CD27-) memory B cells (mBC). D) Percentages of S+RBD+ mBCs of different subsets within S+RBD+ cells pool. Contour plot in A and C are representative from a single subject from each group. Statistical difference was assessed by Kruskal-Wallis test, followed by Dunn’s post-test for multiple comparisons; *P≤ 0.05; **P ≤ 0.01. E) t-SNE visualization of B cells and antigen specific B cells in PLWHIV and healthy donors. t-SNE dimensionality reduction was used to visualize in a bidimensional space a multiparametric dataset. Major B cell subsets (switched memory, swM; unswitched memory uswM; double negative, DN; and naïve) from people living with HIV (PLWHIV; left) and healthy donors (right) were displayed as contour plots in t-SNE map according to the expression of all analyzed markers. Antigen specific B cells (S+/RBS+) were highlighted as dots.

S^+^RBD^+^ B cells were dissected again to identify different populations of memory B cells according to IgD and CD27 expression (Figure 3C). Ig-switched (IgD-/CD27+) and unswitched memory B cells (IgD+/CD27+) were recorded at a significant lower rate in PLWHIV compared to HCs (28.09 ± 20.50% vs 47.32 ± 19.35% and 2.608 ± 7.067% vs 3.509 ± 4.514%; respectively P≤0.05), indicating a limited induction of these populations or a reduced survival through time (Figure 3D). On the contrary, the double negative memory B cells (IgD-/CD27-), also known as atypical or exhausted memory, represented a significantly higher population in PLWHIV compared to HCs (58.90 ± 31.26% vs 41.17 ± 19.07%; P≤0.05). The phenotypic distribution was confirmed by the t-SNE dimensionality reduction analysis (Figure 3 E), which showed a higher frequency of double negative memory B cells in PLWHIV, while switched and unswitched memory were more abundant in HCs. A higher number of spike-specific B cells (dark spot) felled in double negative region in PLWHIV compared with healthy controls where the switched memory phenotype is predominant.

## Discussion

In the present study, we characterized the antibody and memory B cell responses in PLWHIV on ART vaccinated with two doses of a SARS-CoV-2 mRNA vaccine in the 6 months following vaccination and we compared them to HCs.

Almost all PLWHIV developed significant plasma IgG titers after the first administration that significantly increased upon the second dose, remarking the immunogenicity of mRNA vaccines for these patients. Interestingly, anti-spike plasma IgG titers were still significant after 6 months post priming compared to baseline in most patients. Differently from what observed by Portillo and colleagues for anti-RBD antibodies, we did not find any statistical difference when comparing PLWHIV antibody titres to the HCs at all time-points, though it could be influenced by the different assay technology or different mRNA formulations (17). More than 80% of the percentage of PLWHIV showed a positive ACE2/RBD inhibition activity at +60 v2, that declined up to 54% at +150 v2. The reduced performance of neutralizing antibodies in HIV-infected patients, also compared to HC, can theoretically reflect their chronic immune impairment and may be a hint of reduced immune persistence.

A critical point in the persistence of vaccinal immunity overtime is the induction of cellular immune memory. Recent works have demonstrated the presence of spike-specific T cells in PLWHIV vaccinated with two doses of mRNA vaccine, no different compared to HCs (9,24). Here, we demonstrated that spike-specific B cells were at a comparable rate in PLWHIV and HCs. Among these cells, we identified the Ig-switched (IgD-/CD27+), and the unswitched (IgD+/CD27+) memory cells, that maintain IgM and IgD expression. These two populations of spike-specific B memory cells have been observed in SARS-CoV-2 infected as well as in vaccinated subjects and were associated to fast antibody production at future antigen encounter and long-term protection against severe COVID-19 (13,25,26). In PLWHIV both memory cell subsets were significantly lower compared to HC, probably as a consequence of the immune impairment caused by HIV involving both reduced generation and lifespan of memory B cells (18,27). On the other hand, the rate of spike-specific double negative B (IgD-/CD27-) memory cells was significantly higher for PLWHIV. This cell population, also known as terminal memory cells or atypical memory, has been associated to precocious immune ageing and reduced functionality of memory cell compartment and has been already reported in case of HIV infection (28). As this is the first time that memory B cells are evaluated in PLWHIV after mRNA COVID-19 vaccine, the clinical effect of impaired B memory is not known yet.

Stratifying patients according to evidence of enhanced morbidity and mortality for COVID-19 in PLWHIV with low CD4+ T cell count (29), we found that anti-SARS-CoV-2 IgG response of people in the Low CD4+ T cell (≤350/µl) subgroup was generally low, in line to what reported previously (10), maybe reflecting a reduced ability to develop a strong antibody response after the boost. We also found a significant positive correlation, according to Pearson test, between CD4+ T cells/µl and antibody titers at pre v2, +30 v2 and +60 v2. Moreover, when stratified according to CD4+ T cell count no significant differences were observed for mean ACE2/RBD inhibition percentages nor for percentage of neutralization-positive samples. However, the percentage of inhibition positive PLWHIV seemed to be correlated to the CD4+ T cells percentage. Even though the two cohorts of PLWHIV and HCs showed a different sex ratio and BMI, when we analyzed correlates of immunological endpoints in PLWHIV, gender and BMI did not show an association with any outcome. Due to the small number of subjects in the <350 and 350-500 subgroups, we could only make speculations on reduced ability in mounting a humoral immune response. Our cohort represents the general condition of PLWHIV in high income countries, nevertheless a larger study on people with low CD4+ T cells count, uncontrolled HIV and AIDS would be beneficial to describe a more complete scenario.

In summary, our study provides real-world data on immunogenicity of SARS-CoV-2 mRNA vaccines in PLWHIV up to 6 months after vaccination. These results corroborate the opinion that PLWHIV with an adequate CD4+ T cells population (>350 cells/µl) and under ART can mount a significant humoral immune response after vaccination. Interestingly, the longevity of spike-specific humoral immunity in PLWHIV is comparable to healthy controls for the period analysed. Nevertheless, the HIV chronic infection impairs the immune system at different levels and hints of altered immune responsiveness has been observed with ACE2/RBD binding inhibition assay and by spike-specific B cell phenotype.

Findings on other immunocompromised subjects suggest that a third dose can induce a strong immune response even in patients who did not develop immunity after the second dose (30,31). New studies are required to elucidate memory cell longevity and the effect of vaccine boosting. These data are of critical importance to guide vaccination policies and strategies tailored for PLWHIV.

## Data Availability

Individual participant data that underlie the results reported in this article, after de-identification, are available upon reasonable request to the authors

## Contributions

AC, DM, GP, MF and FM conceived the study. AC, DM, GP, MF, FM and MD wrote the protocol. MD and FM looked after ethical approval procedures. IR, BR, MF, MT and FM enrolled patients. MF, EM and GP collected and computerized data. GP, JP, SZ and FF processed the samples. JP, AC, GP, SZ, FF and EP carried out the immunological analysis. AC, MF, JP, GP, SL and DM analyzed the data. MF and FM contribute to data interpretation. JP, MF, AC, BR, FM and DM wrote the manuscript. AC, DM, GP, FM and MF supervised the study. DM provided financial support. All the authors edited and approved the final version of the manuscript.

## Competing interests

MF received speakers’ honoraria, support for travel to meetings, and/or fees for attending advisory boards from Bristol Myers Squibb (BMS), Gilead, Janssen-Cilag, Merck Sharp and Dohme (MSD), and ViiV Healthcare. FM received support for travel to meetings from Angelini, she is principal investigator in sponsor study by TLS (Toscana Life Science) and by GSK Vaccine SRL and she is the contact person for a service contract between GSK Vaccine SRL and Department of Medical Biotechnologies, University of Siena, without receiving any personal remuneration. The other authors declare no conflict of interest regarding this study.

## Funding

This study was supported by the Department of Medical Biotechnologies of the University of Siena (D.M.)

## Data availability

Individual participant data that underlie the results reported in this article, after de-identification, are available upon request.

## Acknowledgments

We would like to thank all the volunteers who participated to the study, the Infectious Diseases Unit Nursing staff who chose to cooperate for blood withdrawal. We thank Giorgio Montesi for data management.

## Notes

### Author Declarations

Ethical Committee for Clinical experimentation of Regione Toscana Area Vasta Sud Est (CEASVE) gave ethical approval for this work

