## Supplementary material for "B cell response six months after SARS-CoV-2 mRNA vaccination in people living with HIV under antiretroviral therapy"

**Table of contents**

**Supplementary Table 1.** Baseline characteristics of people living with HIV (PLWHIV) and healthy controls (HCs). ... 2

**Supplementary Table 2.** Variables associated to ACE2/RBD inhibition percentage at 2 months post second dose (+60 v2) in PLWHIV (univariable and multivariable linear regression analysis). ... 3

**Supplementary Figure 1.** Spike-specific antibody response stratified according to CD4+/µl cells count. **S**tatistical differences between groups at each time point. ... 3

**Supplementary Table 1.** Baseline characteristics of people living with HIV (PLWHIV) and healthy controls (HCs).

| | PLWH N=84 | HC N=79 | p |
|---|---|---|---|
| **General variables** | | | |
| Age, years | 52 (46-58) | 52 (45-60) | 0.732 |
| Male gender | 64 (76.2) | 22 (27.8) | **<0.001** |
| Type of vaccine: | | | |
| -mRNA-1273 | 43 (51.2) | 10 (12.7) | |
| -BNT162b2 | 41 (48.8) | 69 (87.3) | **<0.001** |
| BMI, Kg/m$^2$ | 25.1 (22.9-29.7) | 23.7 (20.8-26.8) | **0.037** |
| **HIV-related variables** | | | |
| Risk factor for HIV infection: | | | |
| -Heterosexual | 14 (16.7) | - | |
| -MSM | 34 (40.5) | - | |
| -IDU | 10 (11.9) | - | |
| -Other/unknown | 26 (31.0) | - | |
| Years from HIV infection | 10.5 (6.3-24.8) | - | |
| CDC stage C | 16 (19.0) | - | |
| HBV or HCV coinfection | 21 (25.0) | - | |
| Zenith HIV-RNA, log10 copies/mL | 5.22 (4.66-5.69) | - | |
| CD4 cell count at nadir, cell/mmc | 154 (34-302) | - | |
| Years from first ART | 7.9 (5.3-13.9) | - | |
| Type of ART: | | | |
| -InSTI + 2NRTI | 36 (42.9) | - | |
| -PI + 2NRTI | 6 (7.1) | - | |
| -NNRTI + 2NRTI | 20 (23.8) | - | |
| -Other | 22 (26.2) | - | |
| Baseline HIV-RNA <50 copies/mL | 76 (90.5) | - | |
| Time from last HIV-RNA >50 copies/mL, years | 4.9 (1.6-8.0) | - | |
| CD4 cell count at baseline, cell/mmc | 639 (425-842) | - | |
| - <=350 cell/mmc | 14 (16.7) | - | |
| - 350-500 cell/mmc | 13 (15.5) | - | |
| - >=500 cell/mmc | 57 (67.9) | - | |
| CD4% | 30.8 (23.7-37.8) | - | |
| CD4%>=30% | 47 (56.0) | - | |
| CD4/CD8 ratio | 0.8 (0.53-1.10) | - | |
| CD4/CD8 ratio >=1 | 31 (36.9) | - | |
| OIR | 28 (33.3) | - | |

**Notes:** values are expressed as n (%), except for * median (interquartile range)

**Abbreviations:** ART, antiretroviral therapy; BMI, body mass index; HBV, hepatitis B virus; HCV, hepatitis C virus; IDU, injecting drug users; InSTI, integrase strand transfer inhibitors; MSM, men who have sex with men; NRTI, nucleoside reverse transcriptase inhibitors; NNRTI, non nucleoside reverse transcriptase inhibitors; OIR, optimal immunological recovery; PI, protease inhibitors.

**Supplementary Table 2.** Variables associated to ACE2/RBD inhibition percentage at 2 months post second dose (+60 v2) in PLWHIV (univariable and multivariable linear regression analysis).

| | **Univariable analysis** | | **Multivariable analysis** | |
|---|---|---|---|---|
| | **Mean change (95% CI)** | **p** | **Adjusted mean change (95% CI)** | **p** |
| Age, per +10 years | 5.01 (-2.85 / 13.03) | 0.204 | | |
| Male gender | -6.18 (-25.65 / 13.29) | 0.527 | | |
| Type of vaccine (mRNA-1273 vs BNT162b2) | 0.06 (-18.28 / 18.39) | 0.995 | | |
| BMI, Kg/m$^2$ | 0.47 (-1.06 / 2.00) | 0.535 | | |
| IDU | -1.46 (-27.95 / 25.03) | 0.912 | | |
| Years from HIV infection, per +10 years | 0.76 (-7.56 / 9.08) | 0.855 | | |
| CDC stage C | -16.45 (-37.41 / 4.51) | 0.121 | | |
| HBV or HCV coinfection | 3.21 (-17.50 / 23.91) | 0.757 | | |
| Zenith HIV-RNA, per +1 log copies/mL | -2.09 (-17.91 / 13.73) | 0.789 | | |
| CD4 cell count at nadir, per +100 cell/mmc | 0.99 (-4.52 / 6.50) | 0.718 | | |
| Years from first ART, per +10 years | 6.98 (-4.35 / 18.31) | 0.221 | | |
| InSTI + 2NRTI | -6.08 (-23.24 / 11.09) | 0.480 | | |
| Baseline HIV-RNA <50 cp/mL | 14.94 (-16.54 / 46.42) | 0.345 | | |
| Time from last HIV-RNA >50 copies/mL, per +10 years | 20.24 (-6.55 / 47.04) | 0.134 | | |
| CD4 cell count at baseline, per +100 cell/mmc | -0.05 (-2.83 / 2.72) | 0.969 | | |
| CD4 cell count at baseline <=350 cell/mmc | -19.30 (-40.99 / 2.40) | 0.080 | -17.18 (-41.54 / 7.18) | 0.163 |
| CD4%>=30% | 8.08 (-8.81 / 24.97) | 0.341 | -2.18 (-26.54 / 22.18) | 0.858 |
| CD4/CD8 ratio >=1 | 10.34 (-6.81 / 27.49) | 0.232 | 7.07 (-17.24 / 31.39) | 0.561 |

**Abbreviations:** ART, antiretroviral therapy; BMI, body mass index; CI, confidence intervals; HBV, hepatitis B virus; HCV, hepatitis C virus; IDU, injecting drug users; InSTI, integrase strand transfer inhibitors; MSM, men who have sex with men; NRTI, nucleoside reverse transcriptase inhibitors; NNRTI, non nucleoside reverse transcriptase inhibitors; OIR, optimal immunological recovery; PI, protease inhibitors.

**Supplementary Figure 1.** Spike-specific antibody response stratified according to CD4+/µl cells count. **S**tatistical differences between groups at each time point.

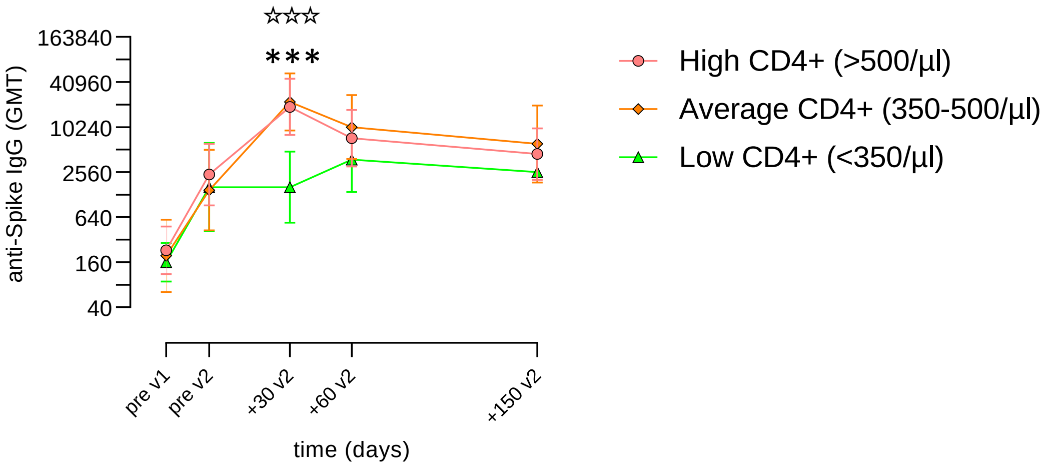


**Legend:** ***P≤ 0.001 (Average CD4+ vs Low CD4+); ☆☆☆P≤ 0.001 (High CD4+ vs Low CD4+).
